# Efficacy and safety of *Boswellia* containing formulations for the treatment of osteoarthritis: protocol for a systematic review and meta-analysis of randomised controlled trials

**DOI:** 10.1101/2020.09.07.20190215

**Authors:** Zhiqiang Wang, Ambrish Singh, Salman Hussain, Pablo Molina García, Benny Antony

## Abstract

Osteoarthritis (OA) is a common chronic joint disease with limited pharmacological management options. *Boswellia* extracts (BE) or formulations containing BE are generally considered as safe and offers a modest efficacy for the treatment of OA. Although previous systematic reviews have assessed the efficacy of BE in OA, these reviews had excluded trials assessing various formulations containing BE and excluded various combinations of BE. Hence, this study aims to systematically review the evidence from RCTs assessing the efficacy and safety of both BE and formulations containing BE for the treatment of OA. Biomedical databases such as PubMed, Embase, and Google Scholar will be searched to identify the RCTs of BE or formulations containing BE in patients with OA (hand, knee, hip, or any other OA). Cochrane risk of bias tool will be used to assess the quality of included studies. Review Manager 5 (Rev Man) and STATA Version 16 will be used to conduct the statistical analysis.

## Background and rationale

Osteoarthritis (OA) is a common chronic joint disease that imparts a substantial and ever-increasing health burden, especially in older people^[1]^. Current treatment guidelines of OA often recommend non-pharmacological treatment as first-line, such as education and self-management, exercise, weight loss if overweight or obese^[2, 3]^. The options for pharmacological management of OA is limited to analgesics, intraarticular corticosteroids, and NSAIDs^[4]^. These medications only have a mild to moderate effect on pain^[5]^, often resulting in patient dissatisfaction and hastening joint replacement^[6]^. There remains unmet healthcare need for chronic pain management^[7, 8]^; as a result, patients are increasingly turning to complementary and alternative medicine^[9]^ such as *Boswellia* extracts (BE) or various formulations containing BE^[10, 11]^.

Frankincense, or BE, has a long history of use (such as religious ceremonies and perfume production), and its medicinal properties have been appreciated for ages^[12]^. Preclinical and clinical evidence suggests that BEs are an effective and safe option for the treatment of OA^[13]^. BEs were often used for rheumatoid arthritis, Crohn’s disease, osteoarthritis, and collagenous colitis^[14]^. Various formulations containing BE often combine BE with other natural products such as curcumin^[15]^, ginger [16], methylsulfonylmethane [17]. A recent systematic review included the randomised clinical trials (RCTs) evaluating the efficacy and safety of BE for the treatment of OA^[18]^. However, studies evaluating formulations containing BE, such as Acujoint™, Lanconone® (E-OA-07), were not included in the review. We hypothesise that BE formulations exhibit a similar or better effect on OA as these combinations often help in increased bio-absorption of the active ingredients of BE such as Acetyl-11-keto-β-boswellic Acid (AKBA).

### Objectives

We aim to systematically review and meta-analyse the evidence from RCTs on the efficacy and safety of BE or formulations containing BE for OA, in terms of pain and physical function improvement, change in biomarkers, and adverse events.

## Methodology

### Eligibility criteria

We will select the eligible RCTs following the specific criteria of PICOS items: Participants, Interventions, Comparators, Outcomes, and Study design. The Preferred Reporting Items for Systematic Reviews and Meta-Analyses (PRISMA) statement will be employed for the conduction and reporting of this systematic review^[19]^. We will include studies published both in English and Chinese.

### Characteristics of participants (P)

We will include adult patients, at least 40 years old, of any sex, and diagnosed with OA according to the criteria proposed by the American College of Rheumatology or similar^[20]^.

### Characteristics of interventions (I)

This study will focus on BE obtained from trees of the genus Boswellia (*Boswellia sacra, B. carterii, B. frereana, B. serrata, and B. papyrifera*) and any formulations containing these BE. RCTs evaluating combination therapy of BE as adjuvant or complementary to conventional drugs (eg. Paracetamol or NSAIDs) will also be included. We will include trials with multi-herbal formulations containing BE and any bioavailability enhancing combination.

### Characteristics of comparators (C)

We will include the studies comparing BE and formulations containing BE with active control (e.g. but not limited to NSAIDs) or placebo for the treatment of OA.

### Characteristics of outcomes (O)

The studies reporting data for at least one of the following outcomes: pain, physical function, imaging biomarkers (x-ray joint space narrowing, MRI structural measures), biochemical markers, medication change, and adverse events will be included in the analysis.

For each outcome, data on baseline and follow-up values and/or mean change from baseline will be extracted. If the data are expressed in the graphical information, the numerical data will be extracted from graphs using the procedure (adapted) suggested by Guyot et al.^[21]^. If the studies do not provide complete data, authors of primary studies will be contacted through email to request missing or additional data.

### Characteristics of study design (S)

Studies will be included if they used a randomised, quasi-randomised, controlled, blinded or non-blinded design. Observational and non-randomised studies will be excluded.

### Primary and secondary outcomes

Our primary outcome will be to evaluate the efficacy of BE and BE formulations on improvement in pain. Furthermore, we will consider improvement in physical function, change in biochemical markers, and imaging biomarkers as secondary outcomes.

## Information sources and search procedure

We will implement the search procedure consistently with the following criteria.

### Electronic source and search strategy

We will retrieve the currently available RCTs on the use of BE and BE formulations for OA through a systematic search of the biomedical database. We will search online databases such as PubMed, Embase, Web of Science, and Cochrane Central Register of Controlled Trial, Google scholar, etc. from inception to September 2020. Search terms for interventions will include: *Boswellia, Boswellia serrata, Boswellia carteri, Boswellia frereana, Boswellia papyrifera, Boswellia sacra*, ru xiang (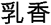, Chinese name for *Boswellia serrata*), boswellic acid, Frankincense, Shallaki, Salai, aflapin, 5-loxin and other formulations such as Acujoint, Lanconone. Search terms for participants will include: Osteoarthritis, Osteoarthritides, Osteoarthrosis, Osteoarthroses, Degenerative Arthritis, Arthritides. Other search terms will be included: randomized controlled trial, random*, random allocation, single-blind, double-blind.

Both published and unpublished trials will be included, with the later including e.g. abstracts, conference proceedings, and posters with available data. Studies that have compared interventions of interest and reported extractable data for at least one measure of pain, physical function, imaging biomarkers, biochemical markers, medication change, and adverse events will be included.

### Hand-searching

Abstract booklet from conference proceedings, abstracts, and poster sessions will be hand searched using the online sources of major international association involved in OA research: European League Against Rheumatism (EULAR), Osteoarthritis Research Society International (OARSI), American Academy of Orthopaedic Surgeons (AAOS) and American College of Rheumatology (ACR). Moreover, references retrieved from relevant meta-analysis and review articles will be hand-searched and analysed for inclusion as per the eligibility criteria.

### Study selection

An Inclusion/Exclusion Form was adapted and used for data extraction (Appendix 1). Study selection will be performed by two reviewers independently, and each potential discrepancy will be discussed and solved through consensus with other authors and independent expert consultation.

### Assessment of risk of bias

The methodological quality of the selected studies will be evaluated using the Cochrane risk of bias tool^[22]^. A Risk-of-Bias form was adapted and used for data extraction (Appendix 2). Each article will be evaluated independently by two researchers using the Cochrane risk of bias tool. The Cochrane risk of bias tool considers characteristics of the following items: sequence generation, allocation concealment, blinding of participants, study personnel and outcome assessors, incomplete outcome data, selective outcome reporting and other potential sources of bias. At the end of quality assessment, a consensus on final evaluation will be reached; any disagreements will be resolved by the discussion with senior authors.

### Data extraction

A data extraction form was adapted and used for data extraction (Appendix 3). Two reviewers will extract data independently from the included studies for the following information: study design, characteristics of the population (age, sex, and BMI), sample size, intervention details and dosage, duration of follow-up, type of placebo or other control, outcome measurements, mean change values of the relevant outcome, and the number of adverse event report and medication change. Intention-to-treat data will be used whenever available. The missing data in the literature will be obtained by emailing the corresponding author; otherwise, it will be estimated by the appropriate method according to Cochrane Handbook 5.1.0^[23]^.

## Statistical analysis

The fixed-effect model will be used if included studies are homogeneous; otherwise, the random-effect model with a restricted maximum-likelihood will be employed for the meta-analysis of both continuous and binary outcomes. The heterogeneity of the effect size across the trials will be tested using the Q statistics (P< 0.05 was considered heterogeneous) and I^2^ statistic (I^2^ > 50% will be considered heterogeneous)^[24]^. Publication bias will be assessed visually with funnel plots^[25]^, and the trim-and-fill method was used to estimate the effect of publication bias (if any)^[26]^.

Due to different outcome measures, the change from baseline to follow-up scores will be translated into standard mean differences (SMD) using Hedges’ g effect sizes, as per the data availability. Subgroup analysis will be conducted based on characteristics of RCTs, such as different formulation types, and comparator types. To further explore the potential heterogeneity among the trials, we will perform the meta-regression of the SMDs on covariates^[27]^, such as baseline characteristics of participants (age, gender, BMI), duration, formulation types (BSE or BSE formulations), type of pain measures (VAS vs. WOMAC/KOOS), dosage, study quality (assessed by the RoB), and types of funding (investigator-initiated vs. industry). Statistical analysis will be performed using STATA version 16 (STATA Corp., Texas, USA) and Review Manager 5 (RevMan 5.3) (Copenhagen: The Nordic Cochrane Centre, The Cochrane Collaboration, 2014).

## Data Availability

Not applicable

## Appendix 1 Inclusion/Exclusion Form for Primary Studies

Identification Details
| Author | Year | Journal/ Conference | Source |
| --- | --- | --- | --- |

Please Tick Only One Box Below
| Included | Excluded | Pending* |
| --- | --- | --- |

\* Issue relates to selective reporting – when authors may have taken pain or physical function measurements, but not reported these within the paper. Reviewers should contact the corresponding author for information on possible non-reported pain or physical function & reasons for exclusion from publication. Study should be listed as ‘Pending’ until clarified. If no clarification is received after three attempts, study should then be excluded.

References to Other Trials
| Did this report include any references to **published reports** of potentially eligible trials not already identified for this review? |  |  |
| --- | --- | --- |
| First author | Journal / Conference | Year of publication |

Did this report include any references to **unpublished data** from potentially eligible trials not already identified for this review? If yes, give list contact name and details

## Appendix 2 Risk-of-Bias Form

Risk-of-Bias Form
| Risk of bias | Author's judgement | Support for judgement |
| --- | --- | --- |
| **Random sequence generation (selection bias)**<br>Was the allocation sequence adequately generated? | Low risk of bias<br>Unclear risk of bias<br>High risk of bias | Describe selection bias (biased allocation to interventions) due to inadequate generation of a randomised sequence |
| **Allocation concealment (selection bias)**<br>Was allocation adequately concealed? | Low risk of bias<br>Unclear risk of bias<br>High risk of bias | Describe selection bias (biased allocation to interventions) due to inadequate concealment of allocations prior to assignment |
| **Blinding (performance bias and detection bias)**<br>Was knowledge of the allocated interventions adequately prevented during the study? | Low risk of bias<br>Unclear risk of bias<br>High risk of bias | Performance bias or detection bias due to knowledge of the allocated interventions after assignment |
| **Incomplete outcome data (attrition bias)**<br>Were incomplete outcome data adequately addressed? | Low risk of bias<br>Unclear risk of bias<br>High risk of bias | Evaluate attrition bias due to amount, nature or handling of incomplete outcome data |
| **Selective reporting (reporting bias)**<br>Are reports of the study free of suggestion of selective outcome reporting? | Low risk of bias<br>Unclear risk of bias<br>High risk of bias | Evaluate reporting bias due to selective outcome reporting |
| **Other bias**<br>Was the study apparently free of other problems that could put it at a high risk of bias? | Low risk of bias<br>Unclear risk of bias<br>High risk of bias | Bias due to problems not covered elsewhere in the table |

## Appendix 3 Data Extraction Form

Study and Participants characteristics
| Study characteristics |  |
| --- | --- |
|  | Further details |
| Country / Countries |  |
| Single centre / multicentre |  |
| Study registration |  |
| Funding |  |
| How was participant eligibility defined?<br>(e.g. How osteoarthritis was defined?) |  |
| How pain was measured? |  |
| How physical function was measured? |  |
| How many people were recruited? |  |
| Duration of study (randomised clinical trial) |  |
| Other |  |
| Participant characteristics |  |
|  | Further details |
| Number of participants (cases versus controls) |  |
| Age (mean, median, range, etc) |  |
| BMI (mean, median, range, etc) |  |
| Female of OA participants (numbers / %, etc) | Female of OA participants (numbers / %, etc) |
| Type of OA (hand, knee, hip, etc) |  |
| Other |  |

| Measures relevant to the review |  |  |  |  |  |
| --- | --- | --- | --- | --- | --- |
| Pain |  |  | VAS / WOMAC/ KOOS |  |  |
| Physical Function |  |  | WOMAC/ KOOS |  |  |
| Adverse events |  |  | YES/ NO |  |  |
| Others |  |  |  |  |  |
| For dichotomous data |  |  |  |  |  |
| OR/ RR<br>(95% CI, p value) | Treatment group (n)<br>n = number of participants, not number of events |  | Control group (n)<br>n = number of participants, not number of events |  |  |
| Other Information (eg. adjustment, cut-off, etc): |  |  |  |  |  |
| For continuous data |  |  |  |  |  |
| Unit | Treatment group |  | Control group |  | Details |
|  | n | Mean (SD) | n | Mean (SD) |  |
| Other relevant data |  |  |  |  |  |

